# Safety and Efficacy of Metformin for Idiopathic Intracranial Hypertension. A U.S-Based Real-World Data Retrospective Multicenter Cohort Study

**DOI:** 10.1101/2024.09.01.24312907

**Authors:** Ahmed Y. Azzam, Mahmoud Nassar, Ahmed Saad Al Zomia, Adam Elswedy, Mahmoud M. Morsy, Adham A. Mohamed, Osman Elamin, Omar S. Elsayed, Mohammed A. Azab, Muhammed Amir Essibayi, Jin Wu, Adam A. Dmytirw, David J. Altschul

## Abstract

**Introduction:** Idiopathic intracranial hypertension (IIH) remains a challenging condition to manage, with limited therapeutic options. This study investigated the potential of metformin as a novel treatment for IIH, exploring its effects on disease outcomes and safety profile.

**Methods:** We conducted a retrospective cohort study using the TriNetX database, analyzing data from 2009 to August 2024. Patients diagnosed with IIH were included, with exclusions for other causes of elevated intracranial pressure and pre-existing diabetes. Propensity score matching was employed to balance cohorts according to age, sex, race, ethnicity, Hemoglobin A1C, and baseline body mass index (BMI) at the time of metformin initiation. Outcomes were assessed at various follow-up points up to 24 months.

**Results:** Our study initially comprised 1,268 patients in the metformin group and 49,262 in the control group, with notable disparities in several parameters. Post-matching, both cohorts were refined to 1,267 patients each after matching with metformin group. Metformin-treated patients showed significantly lower risks of papilledema, headache, and refractory IIH status at all follow-up points (p<0.0001). The metformin group also had reduced rates of therapeutic spinal punctures and acetazolamide continuation. BMI reductions were more pronounced in the metformin group, with significant differences observed from 6 months onward (p<0.0001). Notably, metformin’s beneficial effects persisted independently of BMI changes. The safety profile of metformin was favorable, with no significant differences in adverse events compared to the control group which did not receive metformin during the study timeframe.

**Conclusions:** Our study provides evidence for metformin’s potential as a disease-modifying therapeutic approach in IIH, demonstrating improvements across multiple outcomes. The benefits appear to extend beyond weight loss, suggesting complex mechanisms of action. These findings warrant further investigation through prospective clinical trials to establish metformin’s role in IIH management and explore its underlying therapeutic mechanisms.

## 1. Introduction

The current standard of care for idiopathic intracranial hypertension (IIH) focuses on reducing intracranial pressure (ICP) and preserving visual function [1, 2]. Weight loss remains the cornerstone of therapy, with studies demonstrating significant improvements in ICP and clinical outcomes following a 5-10% reduction in body weight [3, 4]. The Idiopathic Intracranial Hypertension Weight Trial (IIH:WT) provided Class I evidence that bariatric surgery is superior to community weight management programs in reducing ICP and improving quality of life [5]. Pharmacological management primarily involves acetazolamide, a carbonic anhydrase inhibitor that decreases cerebrospinal fluid (CSF) production. The landmark Idiopathic Intracranial Hypertension Treatment Trial (IIHTT) established acetazolamide’s efficacy in improving visual field function and reducing ICP when combined with a low-sodium weight reduction diet [6]. Other therapeutic approaches include topiramate, which offers the dual benefit of ICP reduction and migraine prophylaxis, and surgical interventions such as CSF diversion procedures or optic nerve sheath fenestration for medically refractory cases [7].

Despite these interventions, the management of IIH remains challenging, with a substantial proportion of patients experiencing refractory or recurrent disease [8]. Many patients struggle to achieve or maintain weight loss, particularly through non-surgical means. The side effect profile of acetazolamide, including paresthesia, dysgeusia, and fatigue, often limits its long-term use or dose escalation [9]. Furthermore, a significant proportion of patients experience a plateau in their clinical improvement or require multiple interventions to maintain remission [10]. The lack of targeted therapies addressing the underlying pathophysiology of IIH, particularly the complex interplay between adipose dysfunction, CSF dynamics, and metabolic dysregulation, has hindered progress in disease modification and long-term outcomes [11].

The latest literature evidence has highlighted the unmet need for novel treatment approaches for IIH. Metformin, a biguanide antidiabetic agent, has demonstrated pleiotropic effects beyond glucose control, including modulation of adipose tissue function and reduction of CSF secretion [12]. Preclinical studies have shown that metformin can lower ICP through AMP-activated protein kinase (AMPK)-dependent inhibition of Na+/K+-ATPase at the choroid plexus, suggesting a direct mechanism for CSF production reduction [13]. This effect is particularly intriguing given the recent evidence implicating choroid plexus hypersecretion in IIH pathogenesis [14]. Additionally, metformin’s effects on weight loss, insulin sensitivity, and adipokine profiles may address key pathogenic factors in CSF disorders such as hydrocephalus in rodent models, offering a potential approach to related diseases management in certain phenotypes [15].

The potential of metformin in IIH is further supported by its established safety profile and its ability to mitigate components of metabolic syndrome [16], which are increasingly recognized as contributors to IIH pathophysiology [17]. To address this knowledge gap and explore metformin’s potential as a disease-modifying therapy for IIH, we are conducting a multicenter, retrospective cohort study utilizing the TriNetX database. This large-scale, real-world evidence approach allows for the assessment of metformin’s impact on IIH outcomes across diverse clinical settings in the United States, providing valuable insights into its safety and efficacy in a large patient cohort. Our study aims to evaluate the effects of metformin on IIH-related symptoms, healthcare utilization, and long-term disease progression, offering a robust foundation for future prospective clinical trials. By leveraging this comprehensive dataset, we seek to elucidate metformin’s potential role in expanding the therapeutic armamentarium for IIH, potentially offering a novel, mechanistically targeted approach to this challenging condition.

## 2. Methods

Our study utilized data from the expansive TriNetX Research Network [18], which contains over 200 million electronic health records aggregated from more than 75 healthcare organizations, primarily located in the United States. This comprehensive dataset includes a wide range of patient-level information, such as demographic characteristics, diagnoses, treatments, procedures, and outcomes, all coded using standard medical classification systems like the International Classification of Diseases, 10th Revision (ICD-10) and Current Procedural Terminology (CPT). Researchers can access this extensive real-world data through the secure TriNetX platform to conduct observational studies. Notably, the dataset is regularly updated, ensuring access to the most current and comprehensive healthcare information available. The study protocol was approved by the Institutional Review Board at the Jacobs School of Medicine and Biomedical Sciences, University at Buffalo, NY, USA (STUDY00008628).

We performed a retrospective analysis of the TriNetX data from 2009 to August 2024 (the timeframe associated with individuals with our inclusion and exclusion criteria in TriNetX database), focusing on patients diagnosed with IIH. They excluded individuals with other known causes of elevated intracranial pressure, such as primary brain tumors, secondary brain metastases, cerebral arteriovenous malformations, and venous sinus thrombosis. Patients with pre-existing type 1 or type 2 diabetes mellitus diagnosed before starting metformin or other comparator medications were also excluded. Propensity score matching was used to ensure the study groups were well-balanced in terms of age, sex, race, ethnicity, Hemoglobin A1C, and baseline body mass index (BMI) at the time of metformin initiation. We analyzed the data at different follow-up durations (1-month, 3-months, 6-months, 12-months, and 24-months) and assessed the following outcomes: papilledema, headache, optic atrophy, blindness, pulsatile tinnitus, diplopia, refractory IIH status, visual discomfort, visual field defects, therapeutic spinal puncture rate, and continuation of acetazolamide as the primary treatment.

### 2.1. Statistical Analysis

The TriNetX platform is equipped with a suite of powerful analytical tools, leveraging programming languages such as Java, R, and Python, which enabled the researchers to efficiently query and analyze the comprehensive dataset to extract meaningful insights [18]. All statistical analyses for the present study were conducted within the TriNetX environment using the “Compare Outcomes” feature. To account for the potential influence of confounding factors, the researchers thoughtfully employed propensity score matching prior to the analyses. This involved a 1:1 matching approach, utilizing nearest neighbor matching without replacement and a caliper set at 0.1 times the standard deviation. TriNetX’s proprietary algorithms derive propensity scores through logistic regression, drawing upon matrices of covariates with randomized row order to enhance the robustness of the matching process. The criterion for statistical significance was set at a p-value less than 0.05. This threshold was chosen to balance the need for robust evidence while allowing for the detection of potentially meaningful effects, acknowledging the inherent complexities and nuances present in real-world data [18].

### 2.2. Baseline Demographics

A comprehensive overview of the baseline demographics and clinical characteristics for patients with IIH is presented in **Table 1**, comparing metformin and control groups before and after propensity score matching. Initially, the cohorts comprised 1,268 patients in the metformin group and 49,262 in the control group, with notable disparities in several parameters. Post-matching, both cohorts were refined to 1,267 patients each, achieving remarkable comparability across baseline attributes. The mean age was nearly identical (36.8 vs 37.0 years), with comparable standard deviations. Gender distribution revealed a striking female predominance (93.29% vs 92.66%), consistent with the known epidemiology of IIH. Comorbidity profiles highlighted the complex medical landscape of IIH patients. Endocrine and metabolic diseases were highly prevalent (73.48% vs 73.01%), potentially reflecting the metabolic dysfunction often associated with IIH. Notably, ophthalmological diseases affected approximately 59% of patients in both groups, underscoring the significant ocular manifestations in IIH. Other frequent comorbidities included musculoskeletal diseases, mental and neurodevelopmental disorders, and respiratory conditions, all showing similar distributions between groups.

**Table 1:** Baseline Demographics of The Patients Cohorts.

| Total Patients, n | Before Propensity Score Matching |  | P-Value | After Propensity Score Matching |  | P-Value |
| --- | --- | --- | --- | --- | --- | --- |
|  | Metformin Group | Control Group |  | Metformin Group | Control Group |  |
|  | 1,268 | 49,262 |  | 1,267 | 1,267 |  |
| Mean Age | 36.8 | 36.2 | 0.0323 | 36.8 | 37 | 0.6114 |
| Standard Deviation | 9.66 | 10.1 |  | 9.66 | 10 |  |
| Sex, n (%) |  |  |  |  |  |  |
| Female | 1,182, (93.22%) | 41,006, (83.24%) | < 0.0001 | 1,182, (93.29%) | 1,174, (92.66%) | 0.5340 |
| Male | 53, (4.18%) | 5,503, (11.17%) | < 0.0001 | 53, (4.18%) | 54, (4.26%) | 0.9213 |
| Unknown | 32, (2.52%) | 1,705, (3.46%) | 0.0537 | 32, (2.53%) | 39, (3.08%) | 0.3994 |
| Ethnicity, n (%) |  |  |  |  |  |  |
| Not Hispanic or Latino | 847, (66.80%) | 28,606, (58.07%) | < 0.0001 | 847, (66.85%) | 859, (67.80%) | 0.6113 |
| Unknown Ethnicity | 291, (22.95%) | 15,479, (31.42%) | < 0.0001 | 291, (22.97%) | 282, (22.26%) | 0.6691 |
| Hispanic or Latino | 129, (10.17%) | 4,111, (8.35%) | 0.0378 | 129, (10.18%) | 126, (9.94%) | 0.8430 |
| Race, n (%) |  |  |  |  |  |  |
| White | 730, (57.57%) | 26,556, (53.91%) | 0.0731 | 730, (57.62%) | 726, (57.30%) | 0.8723 |
| Unknown Race | 226, (17.82%) | 10,492, (21.30%) | < 0.0001 | 226, (17.84%) | 223, (17.60%) | 0.6691 |
| Black or African American | 232, (18.30%) | 7,905, (16.05%) | 0.0694 | 232, (18.31%) | 246, (19.42%) | 0.4771 |
| Other Race | 54, (4.26%) | 2,271, (4.61%) | 0.4567 | 54, (4.26%) | 47, (3.71%) | 0.4772 |
| Asian | 18, (1.42%) | 712, (1.45%) | 0.8702 | 18, (1.42%) | 23, (1.82%) | 0.4311 |
| American Indian or Alaska Native | 10, (0.79%) | 161, (0.33%) | 0.0064 | 10, (0.79%) | 10, (0.79%) | 0.9999 |
| Native Hawaiian or Other Pacific Islander | 10, (0.79%) | 117, (0.24%) | 0.0001 | 10, (0.79%) | 0, (0.00%) | 0.0015 |
| Comorbid Diseases, n (%) |  |  |  |  |  |  |
| Endocrine and Metabolic Diseases | 931, (73.42%) | 15,748, (31.97%) | < 0.0001 | 931, (73.48%) | 925, (73.01%) | 0.7877 |
| Ophthalmological Diseases | 750, (59.15%) | 20,859, (42.34%) | < 0.0001 | 750, (59.19%) | 754, (59.51%) | 0.8715 |
| *Musculoskeletal Diseases* | 677, (53.39%) | 13,730,<br>(27.87%) | <<br>0.0001 | 677,<br>(53.43%) | 658,<br>(51.93%) | 0.4497 |
| *Mental and Neurodevelopmental Disorders* | 664, (52.37%) | 13,249,<br>(26.89%) | <<br>0.0001 | 664,<br>(52.41%) | 650,<br>(51.30%) | 0.5778 |
| *Respiratory Diseases* | 581, (45.82%) | 12,811,<br>(26.01%) | <<br>0.0001 | 581,<br>(45.86%) | 581,<br>(45.86%) | 0.9999 |
| *Genitourinary Diseases* | 606, (47.79%) | 10,356,<br>(21.02%) | <<br>0.0001 | 606,<br>(47.83%) | 614,<br>(48.46%) | 0.7504 |
| *Digestive Tract Diseases* | 519, (40.93%) | 10,079,<br>(20.46%) | <<br>0.0001 | 519,<br>(40.96%) | 514,<br>(40.57%) | 0.8398 |
| *Presence of Active Infections* | 386, (30.44%) | 6,770,<br>(13.74%) | <<br>0.0001 | 386,<br>(30.47%) | 375,<br>(29.60%) | 0.6336 |
| *Skin and Subcutaneous Diseases* | 480, (37.85%) | 6,729,<br>(13.66%) | <<br>0.0001 | 480,<br>(37.88%) | 483,<br>(38.12%) | 0.9023 |
| *Circulatory Diseases* | 261, (20.58%) | 5,944,<br>(12.07%) | <<br>0.0001 | 261,<br>(20.60%) | 270,<br>(21.31%) | 0.6604 |
| *Hematological and Immunological Diseases* | 279, (22.00%) | 5,683,<br>(11.54%) | <<br>0.0001 | 279,<br>(22.02%) | 252,<br>(19.89%) | 0.1875 |
| *Active Malignancies (Excluding CNS Tumors and Brain Metastasis)* | 257, (20.27%) | 4,072,<br>(8.27%) | <<br>0.0001 | 257,<br>(20.28%) | 238,<br>(18.78%) | 0.3411 |
| *Presence of Congenital Malformations or Chromosomal Abnormalities* | 112, (8.83%) | 1,955,<br>(3.97%) | <<br>0.0001 | 112,<br>(8.84%) | 105,<br>(8.29%) | 0.6192 |

Abbreviations: CNS: Central Nervous System

## 3. Results

### 3.1. Outcomes Analysis

We performed a longitudinal outcome analysis between the metformin group and the control group in patients with IIH, and the results are presented in **Table 2**. The metformin group consistently demonstrated lower risk percentages for most outcomes compared to the control group. Papilledema, headache, and refractory IIH showed very high statistical significance (p<0.0001) in favor of the metformin group at all follow-up points (1, 3, 6, 12, and 24 months). The risk ratios for these outcomes ranged from 0.238 to 0.889, indicating a substantially lower risk in the metformin group. Optic atrophy risk was similar between the groups at 1, 3, 6, and 12 months, but at 24 months, the metformin group had a slightly higher risk (2.1% vs. 0.8%, p=0.047). Blindness risk was significantly lower in the metformin group at 3 months (p=0.031), but not statistically significant at other follow-up points. Pulsatile tinnitus and diplopia showed significantly lower risks in the metformin group at 6 months (p=0.005 and p=0.007, respectively) and 24 months (p=0.002 and p<0.0001, respectively). However, the differences were not statistically significant at 1, 3, and 12 months. Visual discomfort and visual field defects were significantly lower in the metformin group only at 3 months (p=0.025), with no significant differences at other follow-up durations. The therapeutic spinal puncture rate was significantly lower in the metformin group at all follow-up points (1, 3, 6, 12, and 24 months), with p-values ranging from 0.0001 to 0.007. In addition to that, metformin group achieved greater statistically significant difference in discontinuation of acetazolamide as a mainstay therapy, as the control group had greater higher frequency of continuing acetazolamide during the study timeframe ranging from 6.55% to 21.99%, compared to the control group’s risk ranging from 16.90% to 31.77%. The risk difference and risk ratio favored the metformin group across all durations, with significant p-values (p<0.0001). The 95% confidence intervals for the risk ratios indicated a consistent benefit of metformin over the entire study period.

**Table 2:**
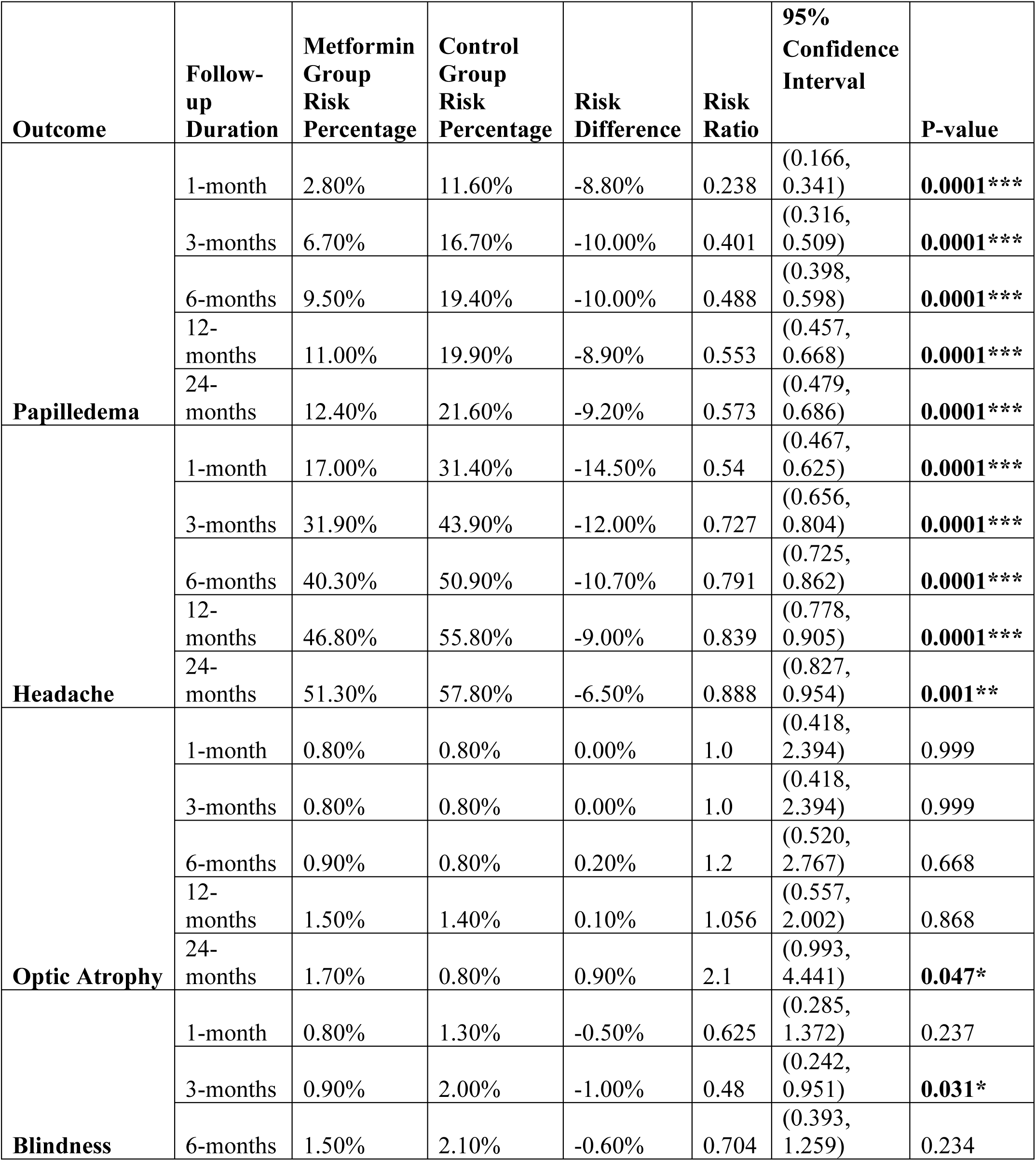
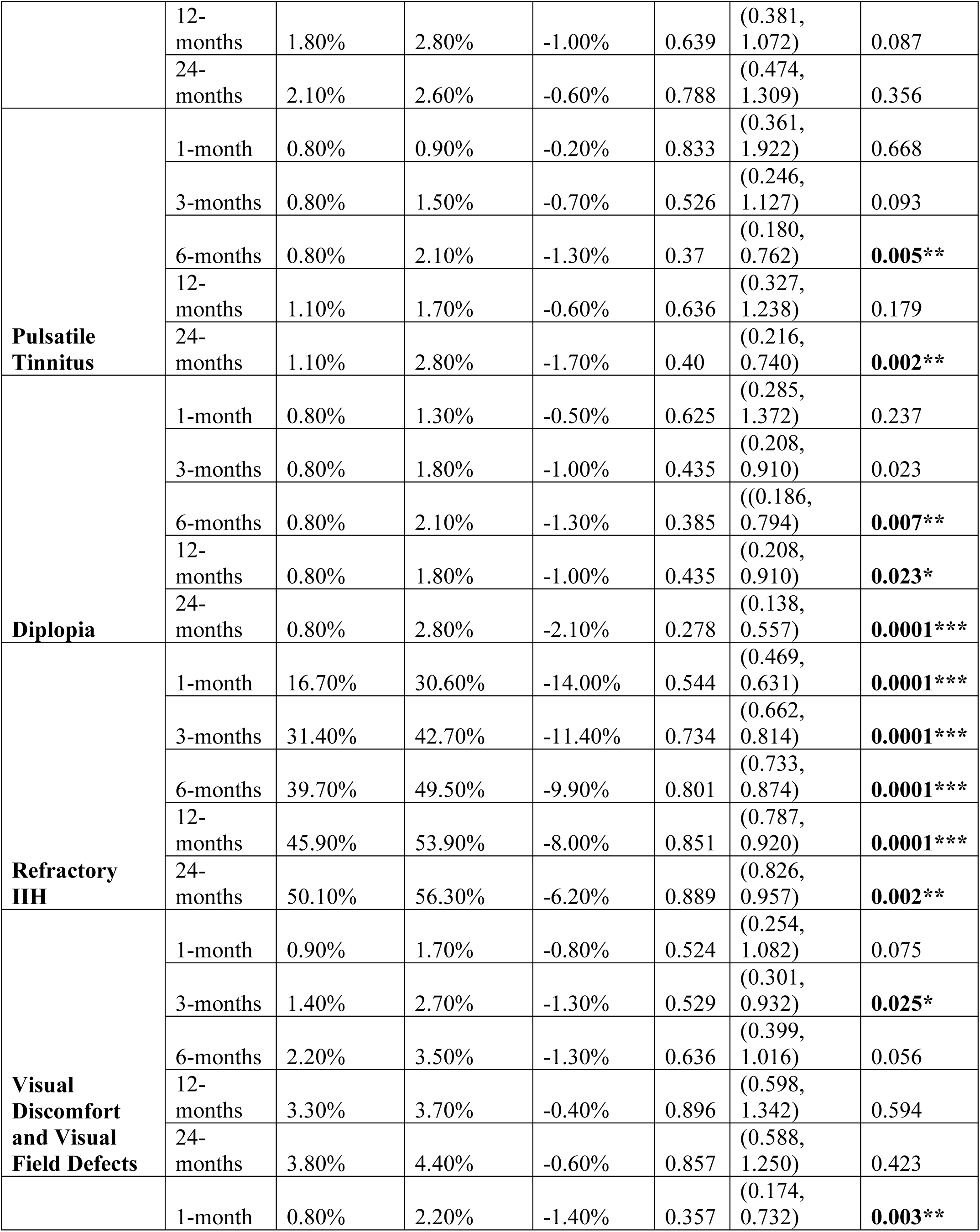
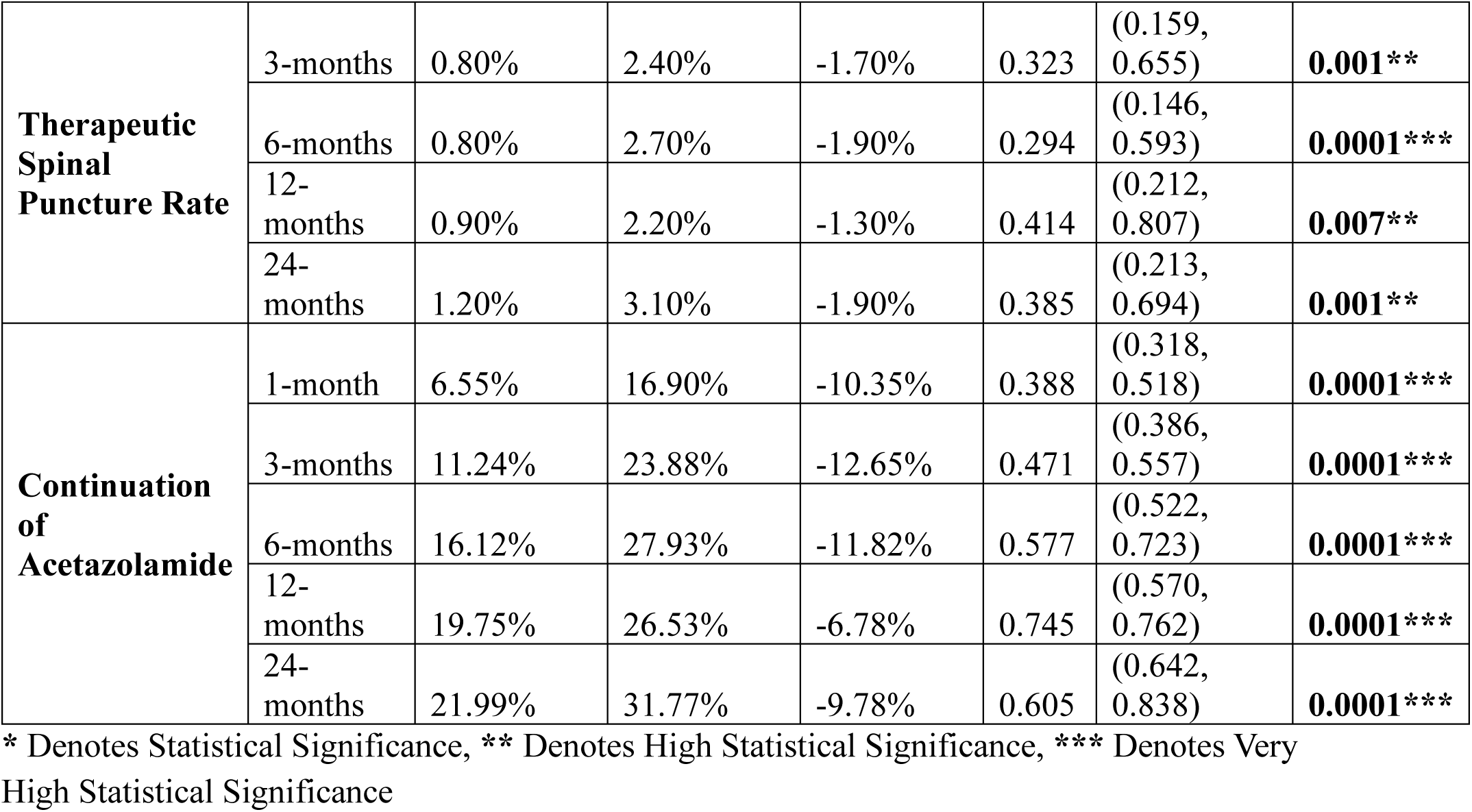
Comparison Between Outcomes and Their Follow-up Duration Between Both Groups Through Propensity Score Matching.

### 3.2. Metformin Dosage Sensitivity Analysis

We performed sensitivity analysis to compare the minimum metformin dose was given to the patients (500mg) compared to all other higher therapeutic doses, as listed in **Table 3**. The sensitivity analysis aimed to determine if the 500mg dose was as effective as higher doses in managing IIH symptoms along different follow-up duration timeframes. The risk percentages for papilledema, headache, optic atrophy, blindness, pulsatile tinnitus, diplopia, refractory IIH, visual discomfort, visual field defects, therapeutic spinal puncture rate, and continuation of acetazolamide were compared between the 500mg dose and all other doses at 1, 3, 6, 12, and 24 months follow-up. The results showed that the risk percentages for all outcomes were similar between the 500mg dose and all other higher doses at each follow-up point. The risk differences between the two groups were minimal, ranging from −1.80% to 0.30%, and none of the differences were statistically significant (p>0.05). These findings suggest that metformin at a dose of 500mg is as effective as higher doses in managing various symptoms associated with IIH. Thes insights are of significant importance, as it indicates that lower doses of metformin may be sufficient to achieve desired therapeutic outcomes, potentially reducing the risk of dose-related adverse effects.

**Table 3:**
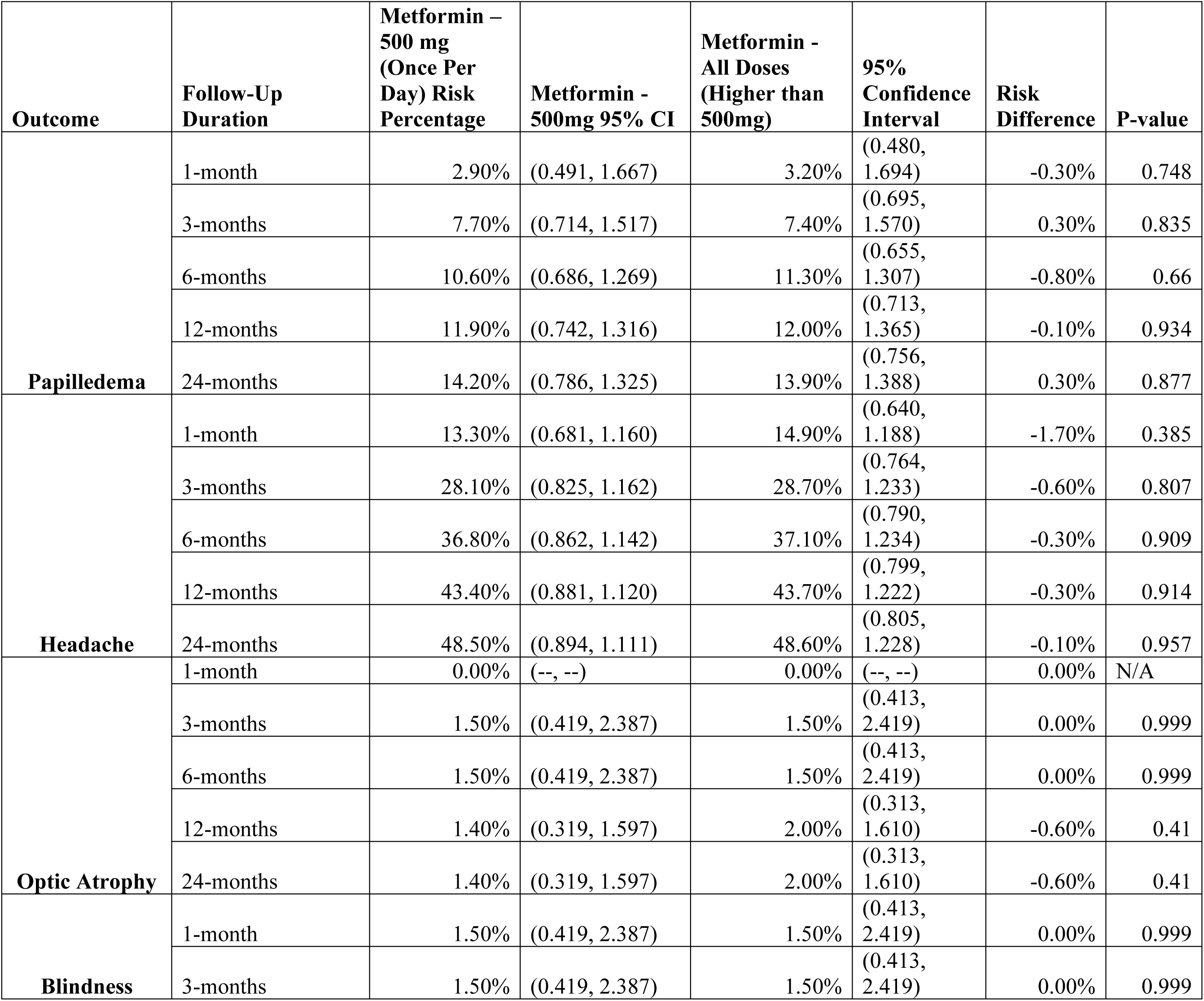
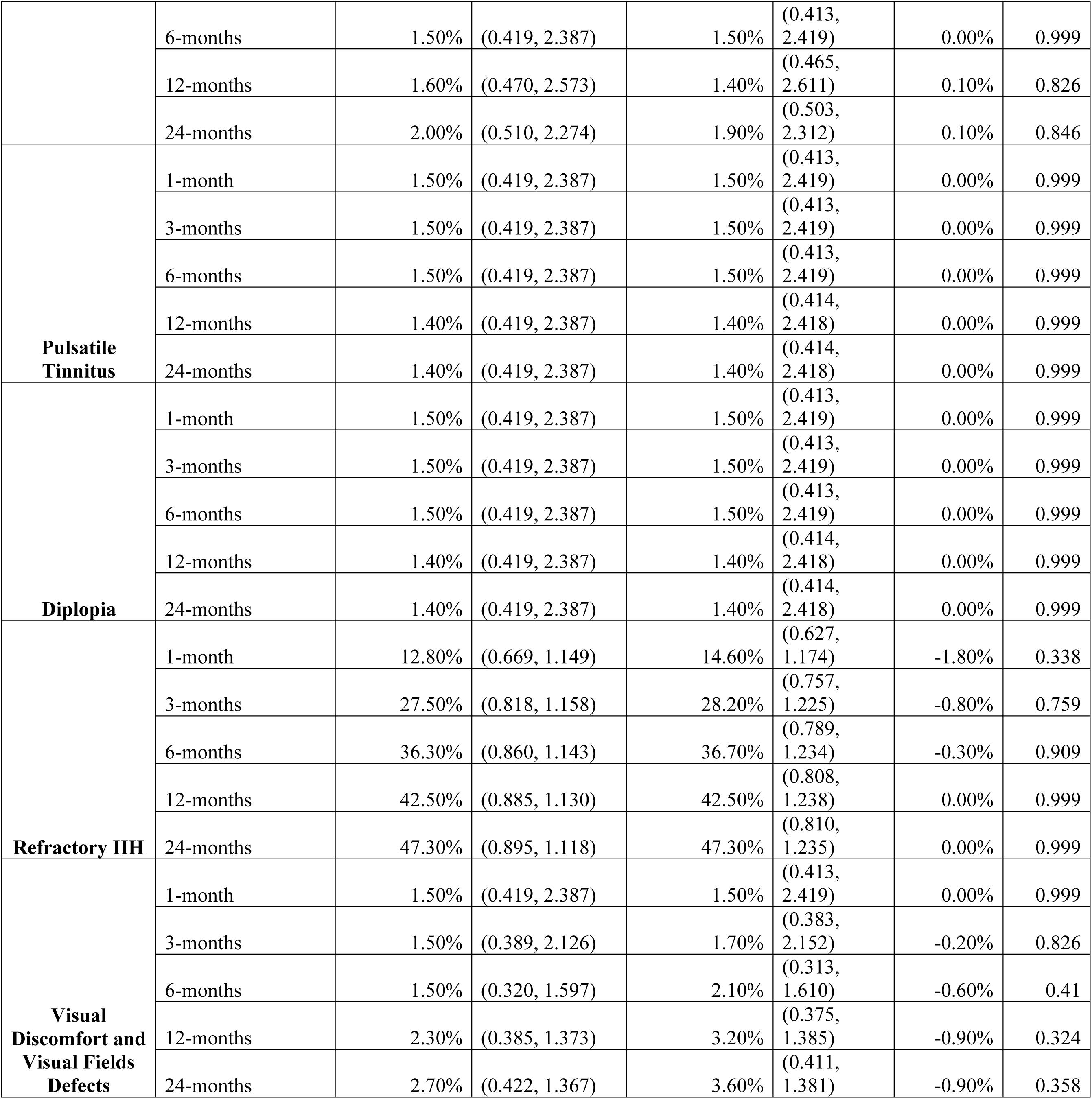
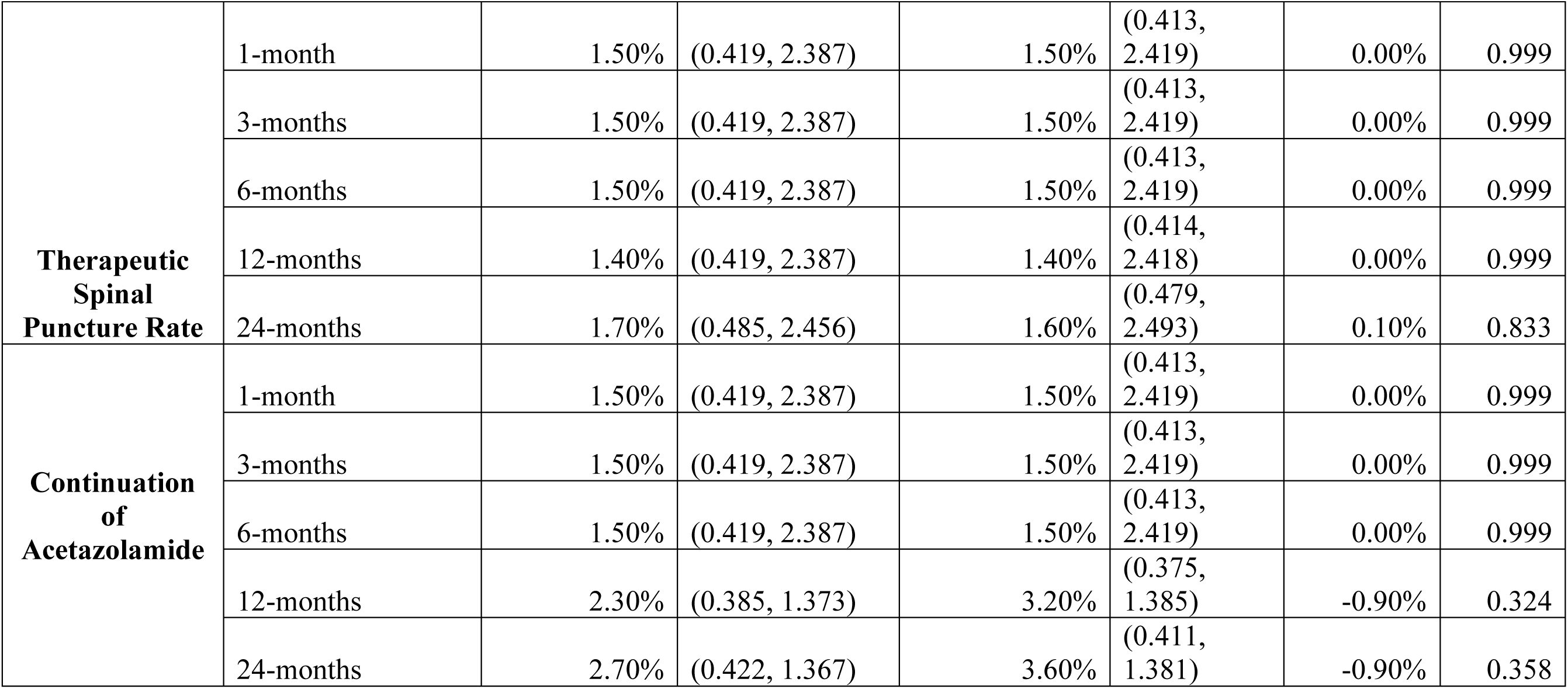
Metformin Dose Sensitivity Analysis, 500mg Compared to All Therapeutic Doses.

### 3.3. Body Mass Index (BMI) Temporal Progression

We analyzed the effects of metformin on BMI over a 24-month period as listed in **Table 4**. Initially, the metformin cohort presented with a notably higher mean BMI (42.03 ± 8.44) than the control group (35.46 ± 8.98). As the study progressed, both groups exhibited BMI reductions, with the metformin group showing more pronounced decreases. Our analysis revealed no statistically significant inter-group differences at one and three months (P = 0.999 and P = 0.1642, respectively). However, from the six-month mark onwards, the metformin group demonstrated significantly greater BMI reductions (P < 0.0001). By the study’s conclusion at 24 months, the mean BMI change difference had reached −1.98 (95% CI, −2.89 to −1.07; P < 0.0001), underscoring the potential long-term efficacy of metformin in weight management for IIH patients. Intriguingly, we performed logistic regression, which included BMI as a factor, indicated that metformin’s beneficial effects on IIH outcomes persisted independently of BMI changes (P-value= 0.891). This finding suggests that metformin may offer additional therapeutic and metabolic benefits beyond weight reduction in IIH management and play a role as a disease-modifying therapeutic option.

**Table 4:** Mean BMI Change Between Metformin and Control Group Along The Follow-up Timeframe.

| Month | Metformin Group |  |  | Control Group |  |  |
| --- | --- | --- | --- | --- | --- | --- |
|  | Sample Size | Mean BMI | SD | Sample Size | Mean BMI | SD |
| 1-month | 288.00 | 42.03 | 8.44 | 381.00 | 35.46 | 8.98 |
| 3-months | 487.00 | 41.53 | 8.44 | 550.00 | 35.72 | 9.06 |
| 6-months | 588.00 | 40.88 | 8.20 | 628.00 | 35.95 | 8.93 |
| 12-months | 664.00 | 40.48 | 8.28 | 691.00 | 35.14 | 8.92 |
| 24-months | 701.00 | 40.41 | 8.55 | 714.00 | 35.82 | 8.96 |
| Mean BMI Change Difference at One Month, (95% CI): | 0.00 (95% CI, -1.33 to 1.33; P= 0.999) |  |  |  |  |  |
| Mean BMI Change Difference at Three Months, (95% CI): | -0.76 (95% CI, -1.83 to 0.31; P= 0.1642) |  |  |  |  |  |
| Mean BMI Change Difference at Six Months, (95% CI): | -1.64 (95% CI, -2.60 to -0.68; P < 0.0001***) |  |  |  |  |  |
| Mean BMI Change Difference at 12 Months, (95% CI): | -1.23 (95% CI, -2.15 to -0.31; P< 0.0001***) |  |  |  |  |  |
| Mean BMI Change Difference at 24 Months, (95% CI): | -1.98 (95% CI, -2.89 to -1.07; P < 0.0001***) |  |  |  |  |  |

\*\*\* Denotes Very High Statistical Significance

### 3.4. Metformin Safety Profile

We analyzed a total of 2,534 patients, equally divided between the metformin and control groups (1,267 patients each) after performing propensity score matching analysis for safety and side effects of metformin. **Table 5** presents a comprehensive overview of the observed side effects, categorized into gastrointestinal, metabolic, and general systemic effects. Our findings indicate that metformin demonstrated a favorable safety profile in IIH patients throughout the study period. Across all categories of side effects, we observed no statistically significant differences between the metformin and control groups (all p-values > 0.05). Gastrointestinal side effects, often associated with metformin use, showed similar incidence rates in both groups. Notably, nausea was reported in 8.52% of metformin users compared to 10.58% in the control group (RR 0.81, 95% CI 0.63-1.03, p=0.09). Vomiting occurred less frequently, affecting 2.37% and 3.31% of the metformin and control groups, respectively (RR 0.71, 95% CI 0.45-1.13, p=0.19). Regarding metabolic side effects, lactic acidosis—a rare but serious concern with metformin use—was observed in 1.03% of metformin users versus 1.74% in the control group (RR 0.59, 95% CI 0.30-1.17, p=0.17). Vitamin B12 deficiency or megaloblastic anemia showed identical rates in both groups (4.58%, RR 1.0, 95% CI 0.70-1.43, p=0.999). General and systemic side effects were also comparable between groups. Myalgia was reported in 6.47% of metformin users and 8.29% of controls (RR 0.78, 95% CI 0.59-1.03, p=0.09), while asthenia affected 5.21% and 5.84% of the metformin and control groups, respectively (RR 0.89, 95% CI 0.65-1.23, p=0.54). These results suggest that metformin is well-tolerated in IIH patients, with a safety profile comparable to that of IIH patients control group who did not undergo metformin therapy during the entire study timeframe.

**Table 5:**
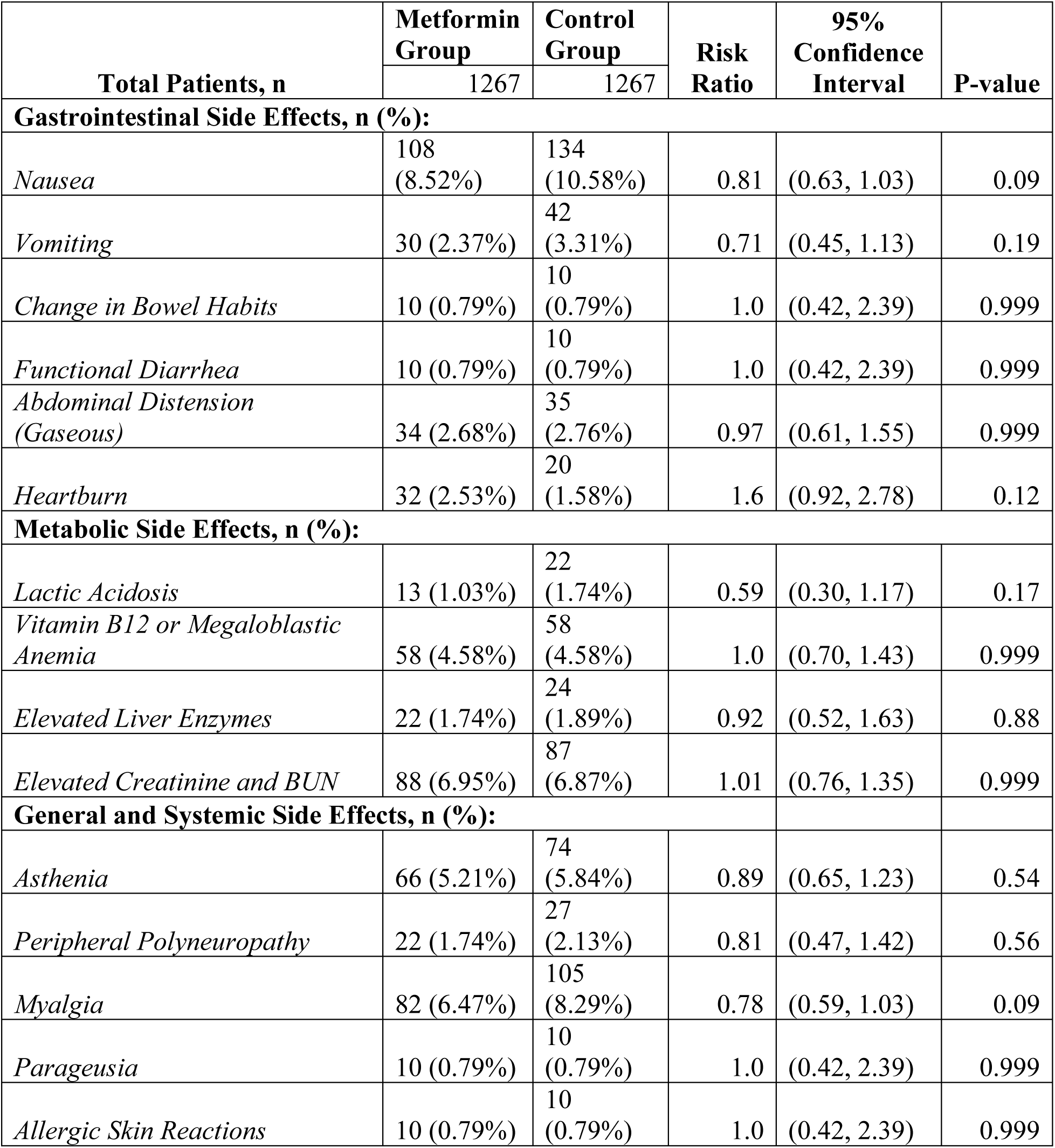
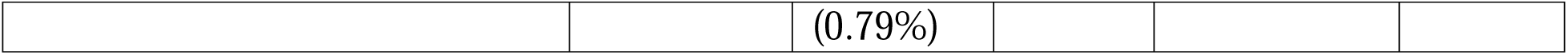
Safety of Metformin Compared to Control Group.

| Total Patients, n | Metformin Group | Control Group | Risk Ratio | 95% Confidence Interval | P-value |
| --- | --- | --- | --- | --- | --- |
|  | 1267 | 1267 |  |  |  |
| Gastrointestinal Side Effects, n (%): |  |  |  |  |  |
| Nausea | 108 (8.52%) | 134 (10.58%) | 0.81 | (0.63, 1.03) | 0.09 |
| Vomiting | 30 (2.37%) | 42 (3.31%) | 0.71 | (0.45, 1.13) | 0.19 |
| Change in Bowel Habits | 10 (0.79%) | 10 (0.79%) | 1.0 | (0.42, 2.39) | 0.999 |
| Functional Diarrhea | 10 (0.79%) | 10 (0.79%) | 1.0 | (0.42, 2.39) | 0.999 |
| Abdominal Distension (Gaseous) | 34 (2.68%) | 35 (2.76%) | 0.97 | (0.61, 1.55) | 0.999 |
| Heartburn | 32 (2.53%) | 20 (1.58%) | 1.6 | (0.92, 2.78) | 0.12 |
| Metabolic Side Effects, n (%): |  |  |  |  |  |
| Lactic Acidosis | 13 (1.03%) | 22 (1.74%) | 0.59 | (0.30, 1.17) | 0.17 |
| Vitamin B12 or Megaloblastic Anemia | 58 (4.58%) | 58 (4.58%) | 1.0 | (0.70, 1.43) | 0.999 |
| Elevated Liver Enzymes | 22 (1.74%) | 24 (1.89%) | 0.92 | (0.52, 1.63) | 0.88 |
| Elevated Creatinine and BUN | 88 (6.95%) | 87 (6.87%) | 1.01 | (0.76, 1.35) | 0.999 |
| General and Systemic Side Effects, n (%): |  |  |  |  |  |
| Asthenia | 66 (5.21%) | 74 (5.84%) | 0.89 | (0.65, 1.23) | 0.54 |
| Peripheral Polyneuropathy | 22 (1.74%) | 27 (2.13%) | 0.81 | (0.47, 1.42) | 0.56 |
| Myalgia | 82 (6.47%) | 105 (8.29%) | 0.78 | (0.59, 1.03) | 0.09 |
| Parageusia | 10 (0.79%) | 10 (0.79%) | 1.0 | (0.42, 2.39) | 0.999 |
| Allergic Skin Reactions | 10 (0.79%) | 10 | 1.0 | (0.42, 2.39) | 0.999 |
|  |  | (0.79%) |  |  |  |

## 4. Discussion

In our large-scale multicenter retrospective study based on TriNetX database, we illustrated compelling evidence for the potential efficacy of metformin as a disease-modifying therapy in IIH. Our findings demonstrate significant improvements across multiple IIH-related outcomes in patients treated with metformin compared to those who did not receive the medication. Notably, the benefits of metformin appear to extend beyond its effects on BMI, suggesting a more complex mechanism of action in IIH pathophysiology.

The marked reduction in papilledema risk observed in the metformin group throughout the study period is particularly striking. This finding aligns with recent research suggesting that metformin may have direct effects on ICP regulation. Botfield et al. [13] demonstrated that metformin can reduce ICP in rodent models of IIH through AMPK-dependent inhibition of the Na+/K+-ATPase at the choroid plexus, thereby decreasing CSF secretion. Our clinical findings support this preclinical evidence, indicating that metformin’s effects on papilledema may be mediated through direct modulation of CSF dynamics rather than solely through weight loss.

The significant reduction in headache prevalence among metformin-treated patients is another key finding of our study. While headache is a cardinal symptom of IIH, its pathophysiology in this context is complex and likely multifactorial. Recent evidence has highlighted the role of calcitonin gene-related peptide (CGRP) in IIH-related headaches, with elevated CGRP levels found in the CSF of IIH patients [19]. Interestingly, metformin has been shown to modulate CGRP signaling in other contexts indirectly, such as attenuating hyperalgesia and allodynia in rodent models with diabetic neuropathy by contributing to the role of AMPK-dependent signaling pathways which play a role in CGRP modulation at the molecular level [20]. This raises the intriguing possibility that metformin’s beneficial effects on IIH-related headaches may involve CGRP modulation and AMPK-dependent regulation pathways, a hypothesis that warrants further investigation.

The observed reduction in refractory IIH status among metformin-treated patients is particularly noteworthy. This finding suggests that metformin may address underlying pathophysiological mechanisms that contribute to treatment resistance in IIH. Recent evidence has implicated adipose tissue dysfunction and altered adipokine profiles in IIH pathogenesis [11]. Metformin’s known effects on adipose tissue function, including modulation of adipokine secretion and improvement of insulin sensitivity, may contribute to its efficacy in refractory cases. Furthermore, emerging evidence suggests that metformin can influence the gut microbiome, which has been increasingly linked to neurological disorders, including those affecting ICP regulation [21]. These multifaceted effects of metformin may explain its potential to improve outcomes in patients who have not responded adequately to conventional therapies.

Our analysis of BMI changes over the study period revealed that while metformin-treated patients experienced greater weight loss, the beneficial effects of metformin on IIH outcomes persisted independently of BMI changes. This finding is crucial as it suggests that metformin’s therapeutic effects in IIH extend beyond weight reduction. Latest evidence has highlighted the importance of metabolic dysfunction in IIH pathogenesis, independent of obesity. For instance, Hornby et al. demonstrated alterations in glucose and lipid metabolism in IIH patients that were not fully explained by BMI [22]. Metformin’s pleiotropic effects on metabolism, including improved insulin sensitivity and modulation of lipid profiles, may therefore contribute to its efficacy in IIH through mechanisms distinct from weight loss.

The potential endocrinological connections underlying metformin’s efficacy in IIH are particularly intriguing. Recent studies have implicated various endocrine factors in IIH pathophysiology, including androgens, glucocorticoids, and growth hormone [23]. Metformin has been shown to influence several of these endocrine pathways. For example, metformin can reduce androgen levels and improve insulin sensitivity in polycystic ovary syndrome (PCOS), a condition often comorbid with IIH [24]. Given that androgen excess has been implicated in IIH pathogenesis, metformin’s androgen-lowering effects may contribute to its therapeutic benefits. Additionally, metformin has been shown to modulate the hypothalamic-pituitary-adrenal (HPA) axis, which could influence CSF dynamics and ICP regulation [25].These endocrinological effects of metformin may explain, in part, its apparent disease-modifying properties in IIH observed in our study.

The safety profile of metformin in our IIH cohort was favorable, with no significant differences in adverse events compared to the control group. This is consistent with metformin’s well-established safety record in other clinical contexts and supports its potential as a long-term therapy for IIH. The similar incidence of lactic acidosis between the metformin and control groups is particularly reassuring, given historical concerns about this rare but serious complication [26].

Our findings have important clinical implications. The observed reductions in papilledema, headache, and refractory disease status suggest that metformin could address multiple aspects of IIH pathophysiology. The potential for metformin to reduce the need for therapeutic spinal punctures and acetazolamide continuation is particularly promising, as it could significantly improve patient quality of life and reduce healthcare utilization. Furthermore, the efficacy of the 500mg dose in our sensitivity analysis suggests that lower doses of metformin may be sufficient to achieve therapeutic benefits in IIH, potentially minimizing dose-related side effects.

While our results are encouraging, several limitations of this study must be acknowledged. The retrospective nature of the analysis introduces potential for confounding and bias, despite our efforts to control for these through propensity score matching. The use of electronic health record data, while providing a large sample size, may be subject to coding errors or incomplete information. Additionally, we were unable to directly measure ICP, lumbar puncture CSF opening pressure, lumbar puncture CSF closing pressure, or perform detailed ophthalmological assessments as these data were not provided for the selected cohort of patients in the research network database, relying instead on documented clinical findings and diagnoses.

Furthermore, while our study suggests a causal relationship between metformin use and improved IIH outcomes, definitive conclusions about causality cannot be drawn from observational data alone. The precise mechanisms by which metformin exerts its effects in IIH remain to be fully elucidated, and our study was not designed to investigate these mechanistic details.

Our large-scale multicenter retrospective study provides compelling evidence for metformin’s potential as a disease-modifying therapy in IIH. The observed benefits, including reduced papilledema risk, decreased headache prevalence, and improved outcomes in refractory cases, suggest that metformin’s effects extend beyond weight loss. These findings align with emerging research on IIH pathophysiology, implicating mechanisms such as CSF dynamics modulation, CGRP signaling, adipose tissue dysfunction, and endocrine factors. The favorable safety profile and potential to reduce invasive interventions further support metformin’s promise as a long-term IIH therapy. While our study has limitations inherent to retrospective analyses, these results provide a strong foundation for future prospective trials to definitively establish metformin’s role in IIH management and elucidate its precise mechanisms of action in this context.

## 5. Conclusions

Our study provides strong evidence for the potential of metformin as a disease-modifying therapy in IIH, with benefits extending beyond weight loss. These findings open new avenues for IIH management and underscore the need for further research into the complex pathophysiology of this condition. Prospective, randomized controlled trials are now warranted to confirm these results and establish optimal treatment protocols. Such studies should include direct measurements of ICP, CSF opening pressure estimations, detailed ophthalmological assessments, and investigations along longitudinal manner into the underlying mechanisms of metformin’s effects in IIH. Additionally, long-term follow-up studies will be crucial to assess the durability of metformin’s benefits and its impact on disease progression. As our understanding of IIH pathophysiology continues to evolve, metformin may represent a promising addition to the therapeutic armamentarium for this challenging condition.

## Acknowledgements

Dr. Wu research is supported by the National Institutes of Health (NIH) under grant agreement, NIH/NINDS grant: R01NS076491.

## IRB Approval

The Institutional Review Board at the Jacobs School of Medicine and Biomedical Sciences, University at Buffalo, NY, USA approved the study protocol under IRB approval number (STUDY00008628).

## Data Availability

Available on TriNetX Database Based on Institutional Collaborations.

## Consent for Participation

N/A

